# NAMPT haploinsufficiency is a therapeutic vulnerability to NAMPT inhibition in -7/-7q MDS

**DOI:** 10.1101/2025.09.14.25335622

**Authors:** Nemo Ikonen, Tanja Ruokoranta, Salla Hyyppä, Ella Sinervuori, Juho J. Miettinen, Joseph Saad, Markus Vähä-Koskela, Caroline A. Heckman

## Abstract

Chromosome 7 abnormalities -7 and -7q define a high-risk subset of myelodysplastic syndromes (MDS) with poor prognosis. The *NAMPT* gene, located at 7q22.3, encodes a rate limiting enzyme (nicotinamide phosphoribosyl transferase) in the NAD+ salvage pathway. Several inhibitors of NAMPT have been developed but their activity in MDS has not been previously described. In this study we investigated if MDS myeloblasts are susceptible to NAMPT inhibition. We show that primary bone marrow cells from patients with -7/-7q MDS exhibit strong and select sensitivity. Bulk viability assays and single cell, multiparametric flow cytometry confirmed enhanced NAMPT inhibitor sensitivity across leukemic cell populations, especially CD34+CD38+ blasts from -7/-7q MDS samples compared to non -7/-7q MDS and healthy donor bone marrow cells. The NAMPT inhibitor KPT-9274 combined with BCL2 inhibitor venetoclax was particularly effective at targeting MDS blasts compared to NAMPT inhibition alone. MDS samples with -7/-7q also showed significantly lower *NAMPT* expression compared to the non -7/-7q samples, indicative of haploinsufficient gene expression profile. In conclusion, these findings support *NAMPT* haploinsufficiency as a vulnerability and as biomarker for NAMPT inhibitor activity in -7/-7q MDS.

## Research letter

Monosomy 7 and deletion of the q arm of chromosome 7 (-7/-7q) are recurrent alterations in myelodysplastic syndromes (MDS) present in approximately 10% of patients, and are associated with poor prognosis, shorter survival, and increased risk of progression to acute myeloid leukemia (AML) (1,2). Nicotinamide phosphoribosyl transferase (NAMPT), encoded by the *NAMPT* gene at chromosome 7q22.3, is the rate limiting enzyme in the nicotinamide adenine dinucleotide (NAD+) salvage pathway (3). NAMPT inhibitors have shown good efficacy in preclinical studies of hematological malignancies as they effectively target cancer cell metabolism by blocking the salvage pathway which leads to NAD+ depletion and cell death (4–7). Previous studies have shown that AML blasts with -7/-7q are exceptionally sensitive to NAMPT inhibitors, with the sensitivity caused by *NAMPT* gene haploinsufficiency as a result of -7/-7q (8,9). The objective of this study was to investigate whether MDS blasts with -7/-7q are similarly susceptible to NAMPT inhibition, and if *NAMPT* haploinsufficiency could be a biomarker for NAMPT inhibitor activity in MDS.

The patient sample cohort is described in **Supplemental Table 1**, and the methods are described in full in the **Supplemental Material** and illustrated in **Figure 1A**. We first assessed the bulk cell viability-based drug sensitivity screening results from bone marrow (BM) cell samples collected from patients with MDS and compared it to data from BM mononuclear cells (MNCs) from healthy donors that were published previously (8,10). MDS and healthy donor BM-MNCs were incubated with the NAMPT inhibitor daporinad for 72 hours, and cell viability was measured using the CellTiter-Glo (CTG) and CellTox Green (CTxG) assays. Dose response curves (**Figure 1B–C**) were summarized as drug sensitivity scores (DSS) **(Figure 1D–E, Supplemental Tables 2–3**). Overall, MDS samples were significantly more sensitive to NAMPT inhibition compared to the healthy donor samples in the CTG assay (**Supplemental Figure 1A**), indicative of an actionable therapeutic window and in line with previous studies in AML (4,5,8,11). The sample carrying -7/-7q was found to be the most responsive sample to NAMPT inhibition (**Figure 1D**). A similar trend was observed in the CTxG assay, with the -7/-7q MDS sample being the most sensitive, but there was no significant difference between MDS and healthy donor samples (**Figure 1E, Supplemental Figure 1B)**. To assess if the -7/-7q MDS samples express less *NAMPT* due to *NAMPT* gene haploinsufficiency, we analyzed the gene expression in CD34+CD38-cells which showed that samples with -7/-7q had significantly lower *NAMPT* expression, indicating a haploinsufficient gene expression profile (**Figure 1F, Supplemental Table 4**).

**Figure 1.**
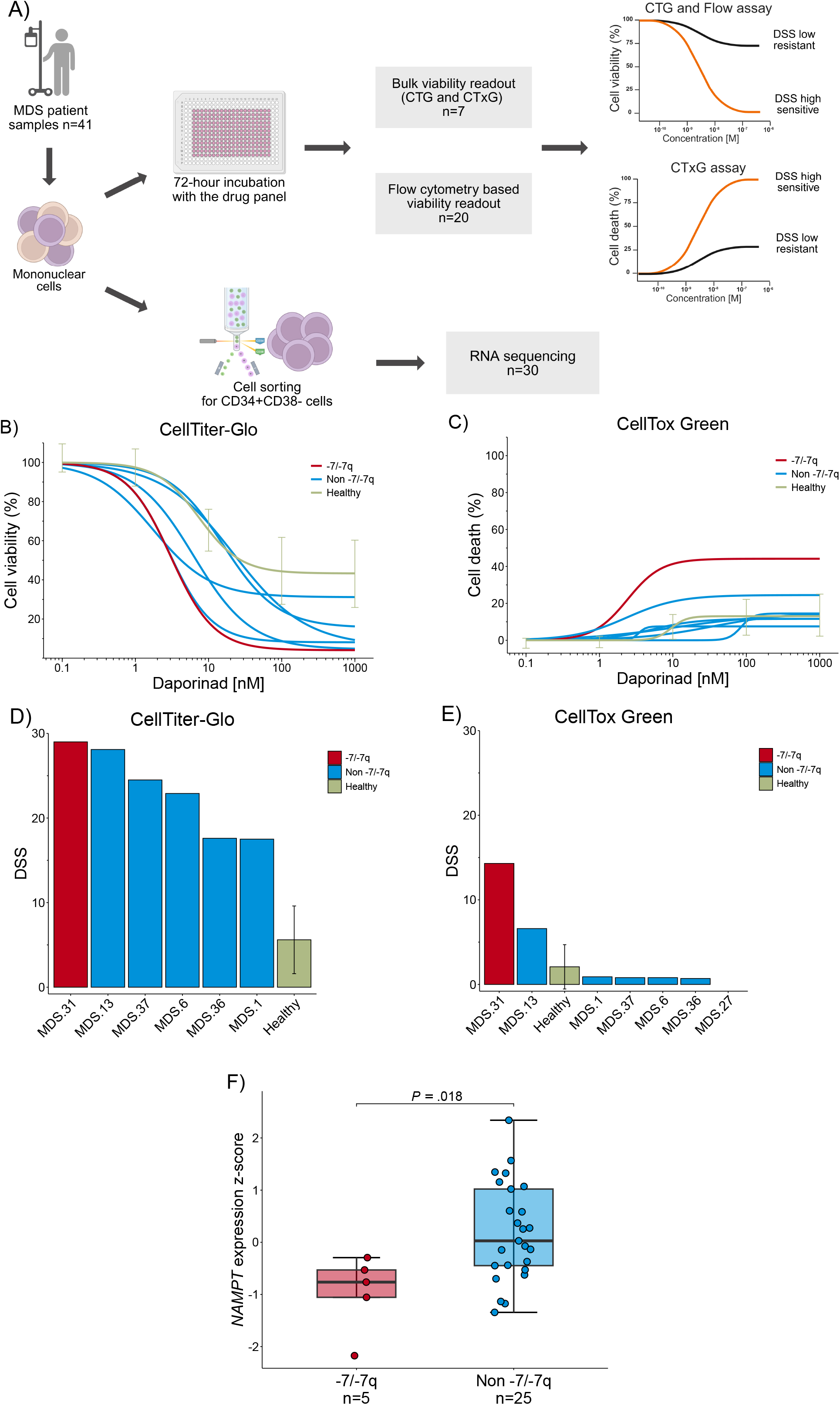
MDS samples are sensitive to NAMPT inhibition. A) Illustration of the experimental workflow. Created with Biorender.com. B) Dose response curves for CellTiter-Glo-based and C) CellTox-Green-based bulk cell viability assays. Samples were treated with daporinad in 5 concentrations for 72 hours. D) Daporinad drug sensitivity score (DSS) values for MDS (n=6) and healthy (n=14) samples for CellTiter-Glo assay. Higher DSS indicates greater sensitivity. Data for healthy samples were summarized as one curve with standard deviations. E) Daporinad DSS values for MDS (n=7) and healthy (n=5) samples for CellTox Green assay. Data for healthy samples (n=5) were summarized as one score with standard deviation. F) *NAMPT* gene expression z-scores were compared between -7/-7q (n=5) and non -7/-7q MDS samples (n=25) using a the two-sample t-test. Gene expression profiles were analyzed from sorted CD34+CD38-cells.

To investigate NAMPT inhibitor activity in different cell subpopulations, we performed flow cytometry-based drug sensitivity analysis. MDS BM-MNCs were incubated with the NAMPT inhibitors daporinad and KPT-9274 for 72 hours (**Supplemental Table 5**) and subsequently stained with an antibody panel (**Supplemental Table 6**). We observed that NAMPT inhibitors effectively target the different myeloblast populations in MDS samples and that CD34+, CD117+, CD34+CD117+, CD34+CD38- and CD34+CD38+ blasts from -7/-7q MDS were significantly more sensitive to NAMPT inhibition by daporinad compared to non -7/-7q MDS blasts (**Figure 2A, Supplemental Figure 2, Supplemental Table 7**). Similar trends were observed for KPT-9274, however, the results were not significant (**Figure 2B**). Additionally, CD34+CD38+ blasts were more sensitive to daporinad and KPT-9274 than CD3+ T lymphocyte cells representing a healthy cell population control and CD14+ monocytic cell population in -7/-7q samples (**Supplemental Figure 3A-B**). These results indicate that the cell populations with a leukemic phenotype, particularly CD34+CD38+, are sensitive to NAMPT inhibition.

**Figure 2.**
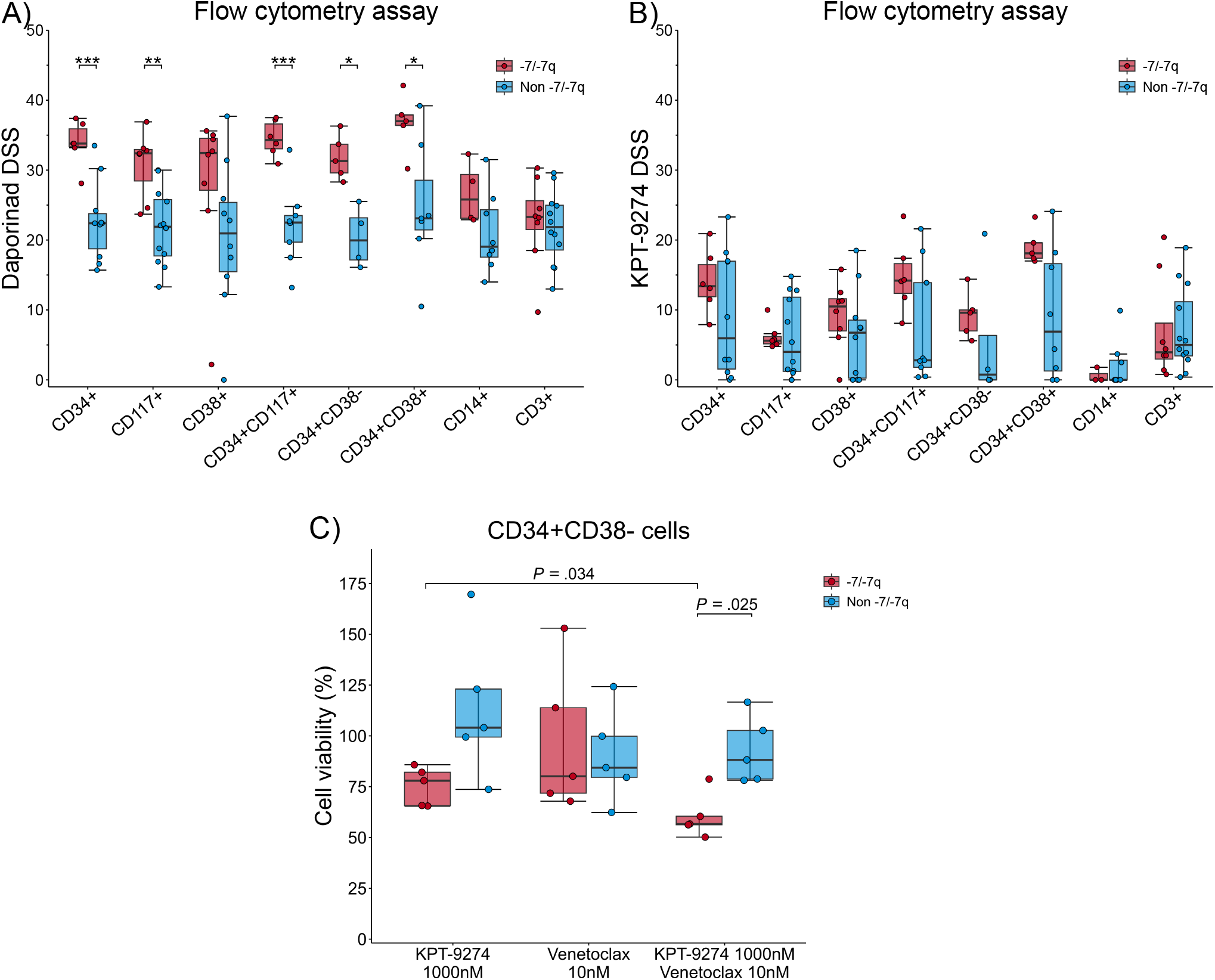
Flow cytometry-based drug sensitivity screening shows that MDS samples with -7/-7q are highly sensitive to NAMPT inhibition. Drug sensitivity scores for the different cell populations for A) daporinad and B) KPT-9274. MDS samples with (n=8) and without (n=12) -7/-7q were incubated with the inhibitors for 72 hours and dose response curves were summarized as DSS values. Higher DSS indicates greater sensitivity. Samples with -7/-7q were compared to those without -7/-7q using a two-sample t-test. C) Comparison of cell viability between single agents and the combination of 1000 nM of KPT-9274 and 10 nM of venetoclax in CD34+CD38-cells in -7/-7q (n=5) and non -7/-7q (n=5) MDS samples. Samples were treated with the inhibitors for 72 hours, after which the cells were stained with detection antibodies to separate different cell types. Comparisons were done using a two-sample t-test and *P* values were adjusted using the Benjamini-Hochberg method.

Next, we assessed KPT-9274 efficacy with three different clinically relevant drug combinations: venetoclax, azacitidine and cytarabine. We observed that the combination of KPT-9274 and venetoclax was significantly more cytotoxic towards CD34+CD38-cells in samples with -7/-7q compared to KPT-9274 alone (**Figure 2C**), and the combination was significantly more cytotoxic in -7/-7q compared to non -7/-7q samples (**Figure 2C**). The addition of azacitidine or cytarabine did not significantly decrease the cell viability (**Supplemental Table 8, Supplemental Figure 3C–D**).

The data from this study demonstrate that NAMPT inhibitors are highly active in -7/-7q MDS *ex vivo* and establish *NAMPT* haploinsufficiency as a vulnerability and biomarker for NAMPT inhibitor activity. The findings suggest that patients with MDS, especially those with -7/-7q, could benefit from NAMPT inhibitors, and support expanded investigations, including clinical evaluation, of NAMPT inhibitors in high-risk MDS.

## Data Availability

The data generated or analyzed during this study are included in this manuscript and its supplementary files. RNA counts data have been deposited to Zenodo: link and DOI will be provided following manuscript acceptance. All other relevant data to the current study are available from the corresponding author (Caroline Heckman) on reasonable request.

## Declarations

### Ethics approval and consent to participate

Bone marrow (BM) or peripheral blood (PB) samples were collected from patients with MDS after informed consent using protocols approved by the Institutional Review Board at the Helsinki University Hospital (permit numbers 239/13/03/00/2010 and 303/13/03/01/2011, Helsinki University Hospital Ethics Committee) in compliance with the Declaration of Helsinki.

### Consent for publication

Not applicable

### Authors’ contributions

N.I., T.R., S.H., E.S., J.M. and J.S. planned and conducted experiments. N.I. analyzed and interpreted the data. N.I., and C.A.H wrote the manuscript. M.V.-K. and C.A.H. provided leadership and funding. All authors read and approved the manuscript.

## Acknowledgements

The authors thank the patients and clinics who made this study possible. The authors also thank the Finnish Hematology Registry and Clinical Biobank (FHRB) for providing the samples and clinical information. Instruments, drug plate preparation and part of the flow cytometry analysis were carried out by the FIMM High Throughput Biomedicine Unit, and library preparation and RNA sequencing were performed by Biomedicum Functional Genomics Unit and FIMM Genomics NGS Sequencing unit, which all are hosted by the University of Helsinki and supported by HiLIFE and Biocenter Finland. Cell sorting and analysis were performed at the HiLife Flow Cytometry Unit, University of Helsinki. N.I. is a Ph.D. candidate at the University of Helsinki. This work is submitted in partial fulfillment of the requirement for the Ph.D. The authors would like to express gratitude to Minna Suvela and Alun Parsons for their role in processing the samples for the study and to Natalia Mokrzecka for her help with the flow cytometry.

